# Effect of Oxygen Saturation Targets on Neurologic Outcomes after Cardiac Arrest: A Secondary Analysis of the PILOT Trial

**DOI:** 10.1101/2025.01.10.25320197

**Authors:** Stephanie C. DeMasi, Alexander T. Clark, Amelia L. Muhs, Jin H. Han, Kipp Shipley, Jared J. McKinney, Li Wang, Todd W. Rice, Ari Moskowitz, Matthew E. Prekker, Nicholas J Johnson, Wesley H. Self, Jonathan D. Casey, Matthew W. Semler, Kevin P. Seitz, Pragmatic Critical Care Research Group

## Abstract

**Background:** More than 600,000 adults in the United States experience a cardiac arrest each year. After resuscitation from cardiac arrest, most patients receive mechanical ventilation. The oxygenation target that optimizes neurologic outcomes after cardiac arrest is uncertain.

**Research Question:** After cardiac arrest, does a lower oxygen saturation (SpO_2_) target improve neurologic outcomes compared to a higher SpO_2_ target?

**Study Design and Methods:** We conducted a secondary analysis of patients who experienced a cardiac arrest before enrollment in the Pragmatic Investigation of optimal Oxygen Targets (PILOT) trial. The PILOT trial assigned critically ill adults receiving mechanical ventilation to a lower (88-92%), intermediate, (92-96%), or higher (96-100%) SpO_2_ target. This subgroup analysis compared patients randomized to a lower-or-intermediate SpO_2_ target (88-96%) versus a higher SpO_2_ target (96-100%) with regard to the primary outcome of survival with a favorable neurologic outcome at hospital discharge (Cerebral Performance Category 1 or 2). The secondary outcome was in-hospital death.

**Results:** Of 2,987 patients in the PILOT trial, 339 (11.3%) experienced a cardiac arrest before enrollment: 221 were assigned to a lower-or-intermediate SpO_2_ target, and 118 were assigned to a higher SpO_2_ target. Overall, the median age was 60 years, 43.5% were female, 58.7% experienced an in-hospital cardiac arrest, and 10.2% had an initial shockable rhythm. Survival with a favorable neurologic outcome occurred in 50 patients (22.6%) assigned to a lower-or-intermediate SpO_2_ target and 15 (12.7%) patients assigned to a higher SpO_2_ target (P=0.03). In-hospital death occurred in 146 patients (66.1%) assigned to a lower-or-intermediate SpO_2_ target and 89 (75.4%) assigned to a higher target (P=0.08).

**Interpretation:** Among patients receiving mechanical ventilation after a cardiac arrest, use of a lower-or-intermediate SpO_2_ target was associated with a higher incidence of a favorable neurologic outcome compared with a higher target. A randomized trial comparing these targets in the cardiac arrest population is needed to confirm these findings.

## INTRODUCTION

More than 600,000 adults in the United States (US) experience a cardiac arrest each year, of whom only 10-25% survive to hospital discharge.^1,2^ Among patients who survive cardiac arrest, brain injury is common, is the leading cause of death, and is a major contributor to long-term disability.^3,4^ After cardiac arrest, nearly all patients receive invasive mechanical ventilation. Clinicians titrate the fraction of inspired oxygen (FiO_2_) on the mechanical ventilator to achieve a target arterial oxygen saturation, which is commonly measured by pulse oximetry (SpO_2_).^3^

After cardiac arrest, patients may be particularly susceptible to ongoing brain injury from extremes of oxygenation. Hypoxemia may worsen the hypoxic-ischemic injury that commonly occurs during the cardiac arrest.^5^ Hyperoxemia may worsen brain injury by inducing cerebral vasoconstriction and creating additional reactive oxygen species.^4^ While preclinical and observational studies have suggested that avoiding hyperoxemia may improve neurologic outcomes in survivors of cardiac arrest,^6–11^ clinical trials evaluating the use of lower oxygenation targets have shown conflicting results, with some suggesting a mortality benefit,^12,13^ some suggesting no effect,^14,15^ and some suggesting harm.^16^ Currently, the International Liaison Committee on Resuscitation guidelines recommend avoiding hyperoxemia with a low certainty of evidence.^5^ Whether use of a lower-or-intermediate SpO_2_ target rather than a higher SpO_2_ target to avoid hyperoxemia prevents further brain injury and increases survival after cardiac arrest remains uncertain.

To address this gap in knowledge, we conducted a secondary analysis of a recent randomized trial that evaluated SpO_2_ targets among critically ill adults receiving invasive mechanical ventilation. We sought to examine the effect of using a lower SpO_2_ target of 88-92% or an intermediate SpO_2_ target of 92-96% compared to a higher SpO_2_ target of 96-100% on outcomes in patients after cardiac arrest. We hypothesized that avoiding hyperoxemia by using a lower-or-intermediate SpO_2_ target would increase the incidence of a favorable neurologic outcome at hospital discharge.

## METHODS

### Study Design

This was a pre-planned subgroup analysis of the Pragmatic Investigation of optimal Oxygen Targets (PILOT) trial, which was a cluster-randomized, cluster-crossover trial in adult patients receiving invasive mechanical ventilation in the medical intensive care unit or the emergency department at a single academic medical center.^17^ The PILOT trial compared the effects of lower, intermediate, and higher SpO_2_ targets on ventilator-free days and in-hospital mortality among critically ill adults receiving mechanical ventilation and has been published previously.^17^ Patients were enrolled at the first receipt of mechanical ventilation in a study unit between July 1, 2018, and August 31, 2021. The PILOT trial was approved by the institutional review board at [redacted for peer review].

### Population

All patients enrolled in the PILOT trial were screened for inclusion. To maximize our sample size, this included patients enrolled during one of the prespecified analytic washout periods in which patients received the assigned SpO_2_ target but were not included in the primary analysis of the trial. Patients who experienced a cardiac arrest (defined as the receipt of chest compressions or defibrillation) with sustained return of spontaneous circulation (defined as spontaneous and sustained return of a pulse for 20 or more consecutive minutes) prior to enrollment were included in the analysis.^18^ The PILOT trial prespecified cardiac arrest as a subgroup that was expected to modify the effect of the intervention on outcomes.^19^

### Study Interventions

The trial protocol instructed respiratory therapists to adjust the FiO_2_ administered in order to achieve an SpO_2_ within the assigned target range from initiation of mechanical ventilation until discontinuation or transfer out of the study unit. For this analysis, we reduced the three trial groups in PILOT (SpO_2_ 88-92%, 92-96%, and 96-100%) to two groups in order to compare the lower-or-intermediate SpO_2_ target group (SpO_2_ 88-96%) versus the higher SpO_2_ target group (SpO_2_ 96-100%). The rationale for combining the lower and intermediate SpO_2_ groups was to facilitate the examination of our hypothesis that avoiding hyperoxemia, which was a goal of both the lower and intermediate groups, would improve outcomes.

### Data Collection

Trial personnel collected data on baseline characteristics and in-hospital outcomes from the electronic medical record. These included data on the initial presenting rhythm (shockable or non-shockable), witnessed (bystander, medical team, or unwitnessed) and cause of arrest (defined as the most likely primary cause of the arrest as determined by physician review and classified according to the Ustein guidelines).^20^ Cardiac arrests from a medical cause were categorized as being due to respiratory failure, cardiac, shock, or other medical causes. Cardiac arrests not from a medical cause included cardiac arrests due to trauma, drug overdose, drowning, or asphyxia.^20^ Data on SpO_2_ and FiO_2_ after enrollment were automatically extracted from the bedside monitor at a frequency of every one minute.^21^

### Study Outcomes

The primary outcome was a favorable neurologic outcome at hospital discharge, defined as a Cerebral Performance Category of 1 (good cerebral performance) or 2 (moderate cerebral disability) at the time of hospital discharge.^22–25^ The Cerebral Performance Category scale is a widely-used and validated measure for assessing the functional status of patients after cardiac arrest, and is predictive of long-term outcomes.^22,23,26^ The scale consists of 5 mutually exclusive categories: 1, good cerebral performance; 2, moderate cerebral disability; 3, severe cerebral disability; 4, coma or vegetative state; 5, death. Data regarding patients’ Cerebral Performance Category at hospital discharge were recorded by two independent study physicians blinded to trial group assignment after independent reviews of the entire electronic medical record using a standardized case report form. A third blinded physician resolved cases with a disagreement between the initial two reviewers. Three-way disagreements were resolved by discussion among the three reviewers until a consensus was reached. This method of electronic medical record review and Cerebral Performance Category determination has been previously reported and validated.^22^

The secondary outcome was in-hospital death from any cause. Exploratory outcomes included the number of days alive and free of invasive mechanical ventilation to 28 days (ventilator-free days)^27^ and the location of discharge, dichotomized as discharge to home (home with or without skilled care) versus all other discharge destinations including death.

### Power Calculation

We calculated that the fixed sample size of 339 patients (221 in the lower-or-intermediate SpO_2_ target group and 118 in the higher target group) would provide 80% statistical power at a two-sided alpha level of 0.05 to detect an absolute difference of greater than 12.0% in a favorable neurological outcome at hospital discharge, assuming an incidence of 11% in the higher target group.^28,29^

#### Statistical Analysis

The primary analysis was an unadjusted, intention-to-treat comparison of the incidence of a favorable neurologic outcome (primary outcome) between patients assigned to a lower-or-intermediate SpO_2_ target versus patients assigned to a higher SpO_2_ target using a chi-squared test.

We performed two additional analyses of Cerebral Performance Category. In the first, we fit a multivariable logistic regression model for the primary outcome with independent variables of trial group assignment and the following prespecified baseline covariates: age, sex, initial shockable rhythm, witnessed (bystander, medical team, or unwitnessed), cause of arrest, and the date of enrollment in the trial. In the second, we analyzed the Cerebral Performance Category as an ordinal outcome ranging from 1 (good cerebral performance) to 5 (death) using a proportional odds model, with an odds ratio (OR) greater than 1.0 indicating a more favorable outcome with a lower-or-intermediate SpO_2_ target compared to a higher SpO_2_ target.

The secondary outcome (in-hospital death) and exploratory outcomes (discharge to home; ventilator-free days) were compared between the two study groups using a chi-squared test for dichotomous outcomes and a proportional odds model for ordinal outcomes. Analyses were performed using STATA version 18.0 and R version 4.1.0 (R Foundation for Statistical Computing).

## RESULTS

Of the 2,987 patients enrolled in the PILOT trial, 378 patients had experienced a cardiac arrest when assessed at enrollment in the trial. Of these, 27 were excluded because the onset of the cardiac arrest was not prior to trial enrollment, 7 were excluded because they did not receive chest compressions or defibrillation, and 5 were excluded because return of spontaneous circulation was not sustained (Figure 1).^18^ The remaining 339 patients were included in this analysis, 221 of whom were assigned to the lower-or-intermediate SpO_2_ targets and 118 of whom were assigned to the higher SpO_2_ target. Overall, the median age was 60 years, 43.5% were female, 58.7% experienced an in-hospital cardiac arrest, and 10.2% had an initial shockable rhythm. The trial groups had similar characteristics at baseline (Table 1).

**Figure 1.**
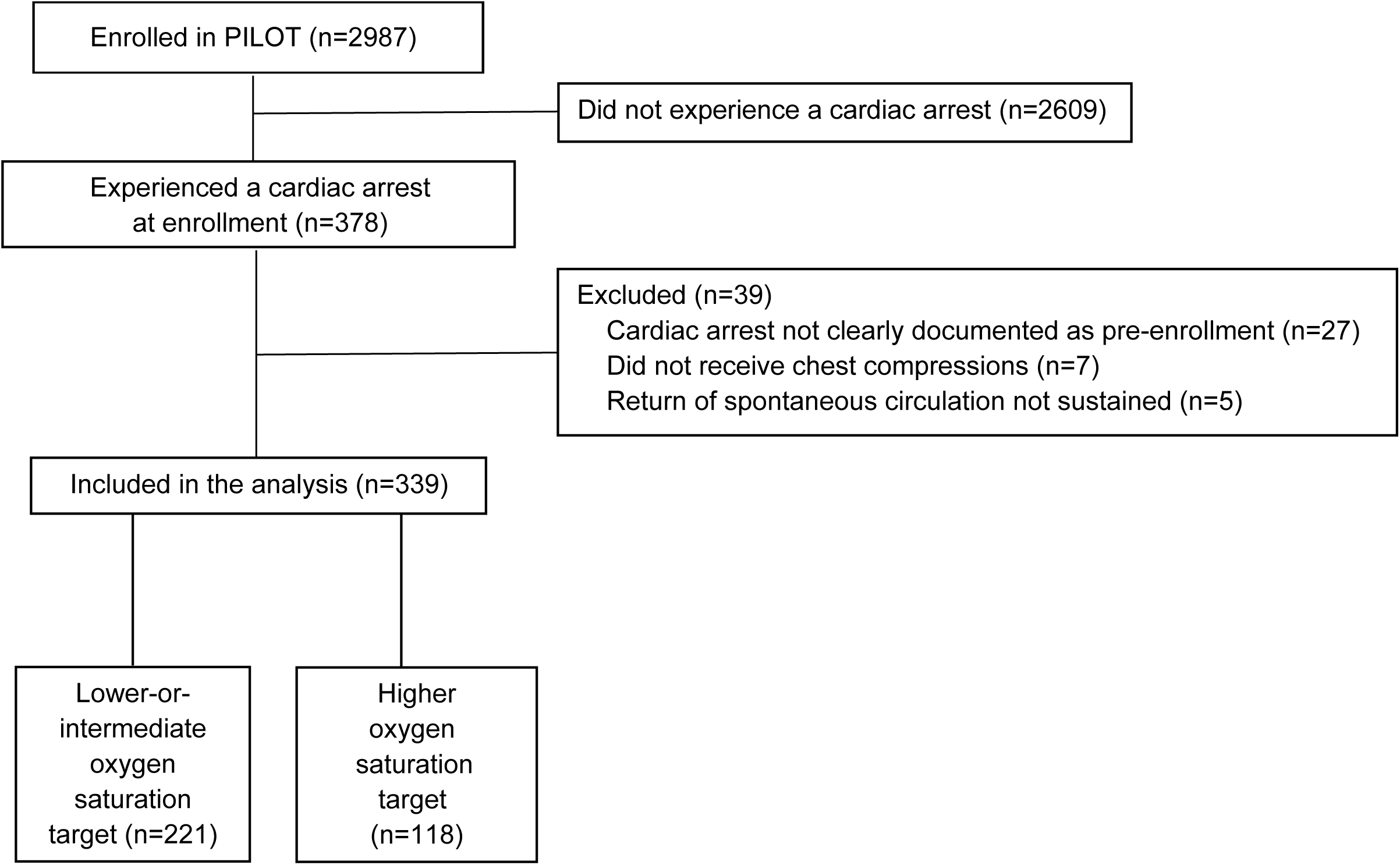
Number of patients screened, excluded, and included in the analysis. Abbreviations: PILOT = *Pragmatic Investigation of Oxygen Targets trial*

**Table 1.**
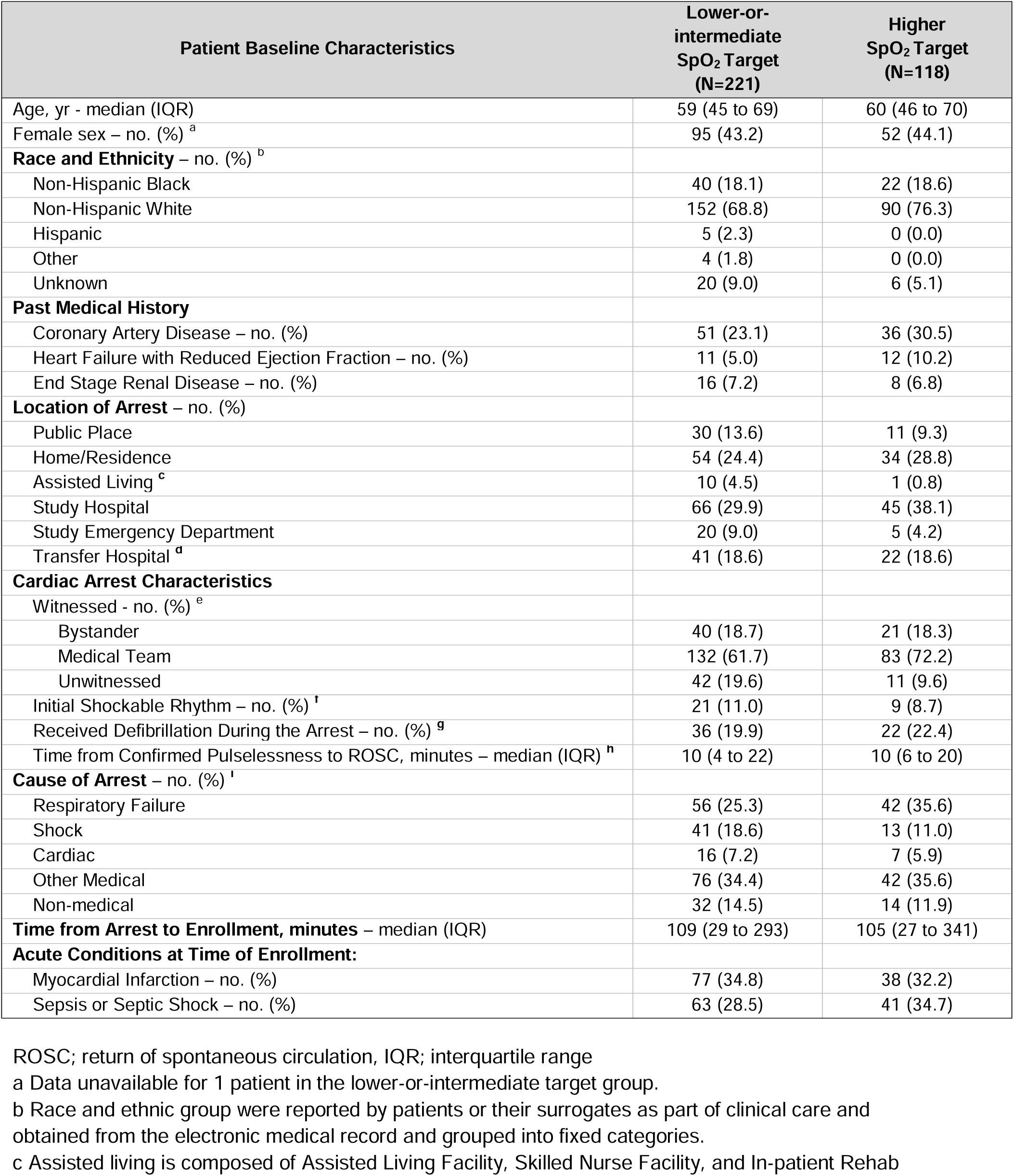
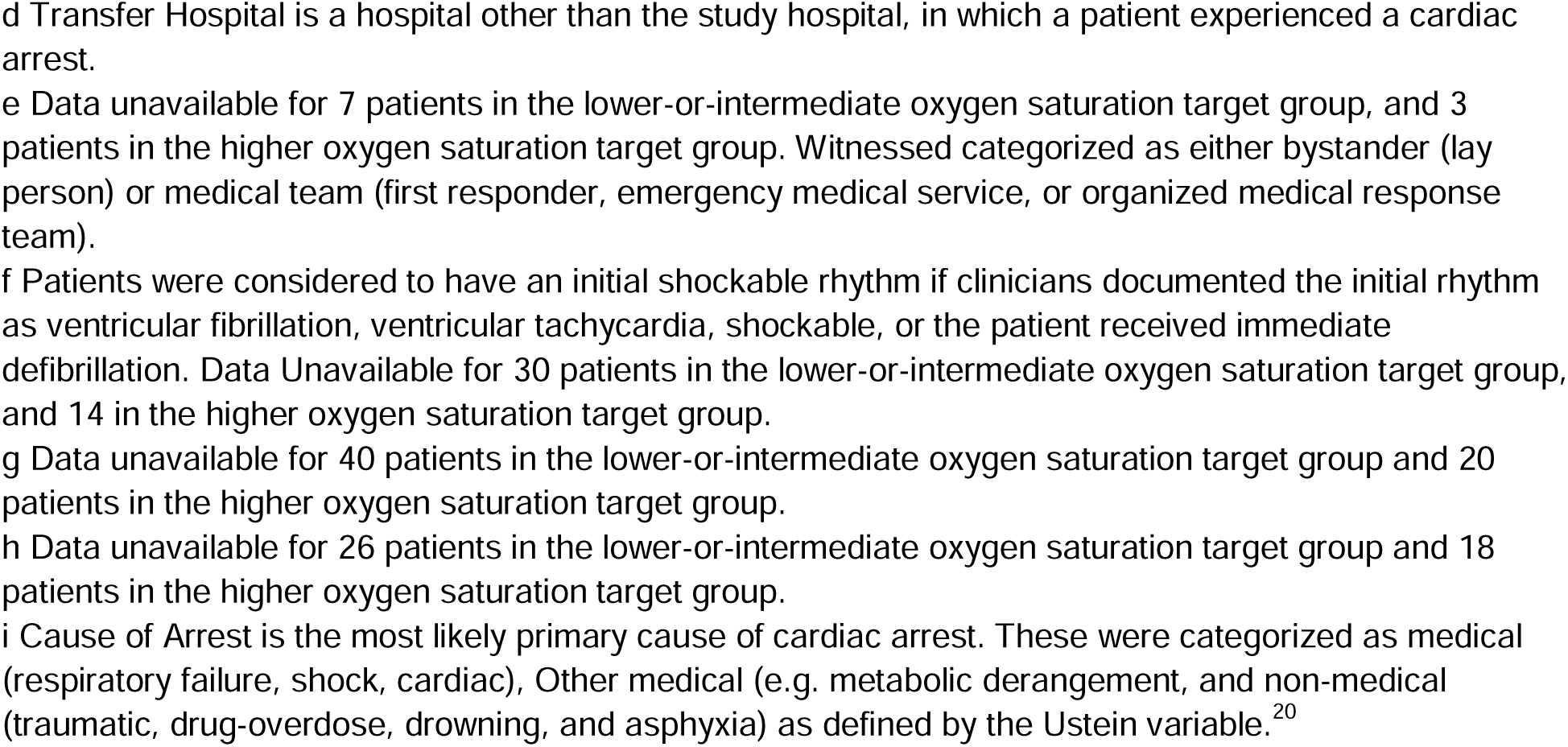
Baseline Characteristics.

| Patient Baseline Characteristics | Lower-or-intermediate<br>SpO<sub>2</sub> Target<br>(N=221) | Higher<br>SpO<sub>2</sub> Target<br>(N=118) |
| --- | --- | --- |
| Age, yr - median (IQR) | 59 (45 to 69) | 60 (46 to 70) |
| Female sex – no. (%) <sup>a</sup> | 95 (43.2) | 52 (44.1) |
| **Race and Ethnicity – no. (%) <sup>b</sup>** |  |  |
| Non-Hispanic Black | 40 (18.1) | 22 (18.6) |
| Non-Hispanic White | 152 (68.8) | 90 (76.3) |
| Hispanic | 5 (2.3) | 0 (0.0) |
| Other | 4 (1.8) | 0 (0.0) |
| Unknown | 20 (9.0) | 6 (5.1) |
| **Past Medical History** |  |  |
| Coronary Artery Disease – no. (%) | 51 (23.1) | 36 (30.5) |
| Heart Failure with Reduced Ejection Fraction – no. (%) | 11 (5.0) | 12 (10.2) |
| End Stage Renal Disease – no. (%) | 16 (7.2) | 8 (6.8) |
| **Location of Arrest – no. (%)** |  |  |
| Public Place | 30 (13.6) | 11 (9.3) |
| Home/Residence | 54 (24.4) | 34 (28.8) |
| Assisted Living <sup>c</sup> | 10 (4.5) | 1 (0.8) |
| Study Hospital | 66 (29.9) | 45 (38.1) |
| Study Emergency Department | 20 (9.0) | 5 (4.2) |
| Transfer Hospital <sup>d</sup> | 41 (18.6) | 22 (18.6) |
| **Cardiac Arrest Characteristics** |  |  |
| Witnessed - no. (%) <sup>e</sup> |  |  |
| Bystander | 40 (18.7) | 21 (18.3) |
| Medical Team | 132 (61.7) | 83 (72.2) |
| Unwitnessed | 42 (19.6) | 11 (9.6) |
| Initial Shockable Rhythm – no. (%) <sup>f</sup> | 21 (11.0) | 9 (8.7) |
| Received Defibrillation During the Arrest – no. (%) <sup>g</sup> | 36 (19.9) | 22 (22.4) |
| Time from Confirmed Pulselessness to ROSC, minutes – median (IQR) <sup>h</sup> | 10 (4 to 22) | 10 (6 to 20) |
| **Cause of Arrest – no. (%) <sup>i</sup>** |  |  |
| Respiratory Failure | 56 (25.3) | 42 (35.6) |
| Shock | 41 (18.6) | 13 (11.0) |
| Cardiac | 16 (7.2) | 7 (5.9) |
| Other Medical | 76 (34.4) | 42 (35.6) |
| Non-medical | 32 (14.5) | 14 (11.9) |
| **Time from Arrest to Enrollment, minutes – median (IQR)** | 109 (29 to 293) | 105 (27 to 341) |
| **Acute Conditions at Time of Enrollment:** |  |  |
| Myocardial Infarction – no. (%) | 77 (34.8) | 38 (32.2) |
| Sepsis or Septic Shock – no. (%) | 63 (28.5) | 41 (34.7) |

ROSC; return of spontaneous circulation, IQR; interquartile range
a Data unavailable for 1 patient in the lower-or-intermediate target group.
b Race and ethnic group were reported by patients or their surrogates as part of clinical care and obtained from the electronic medical record and grouped into fixed categories.
c Assisted living is composed of Assisted Living Facility, Skilled Nurse Facility, and In-patient Rehab
d Transfer Hospital is a hospital other than the study hospital, in which a patient experienced a cardiac arrest.
e Data unavailable for 7 patients in the lower-or-intermediate oxygen saturation target group, and 3 patients in the higher oxygen saturation target group. Witnessed categorized as either bystander (lay person) or medical team (first responder, emergency medical service, or organized medical response team).
f Patients were considered to have an initial shockable rhythm if clinicians documented the initial rhythm as ventricular fibrillation, ventricular tachycardia, shockable, or the patient received immediate defibrillation. Data Unavailable for 30 patients in the lower-or-intermediate oxygen saturation target group, and 14 in the higher oxygen saturation target group.
g Data unavailable for 40 patients in the lower-or-intermediate oxygen saturation target group and 20 patients in the higher oxygen saturation target group.
h Data unavailable for 26 patients in the lower-or-intermediate oxygen saturation target group and 18 patients in the higher oxygen saturation target group.

A total of 864,886 SpO_2_ values were measured between enrollment and cessation of mechanical ventilation among the 339 patients, with a median interval between SpO_2_ measurements of 1 minute. Oxygen saturation values were lower among patients assigned to the lower-or-intermediate SpO_2_ targets than those assigned to the higher SpO_2_ target (Figure 2).

**Figure 2.**
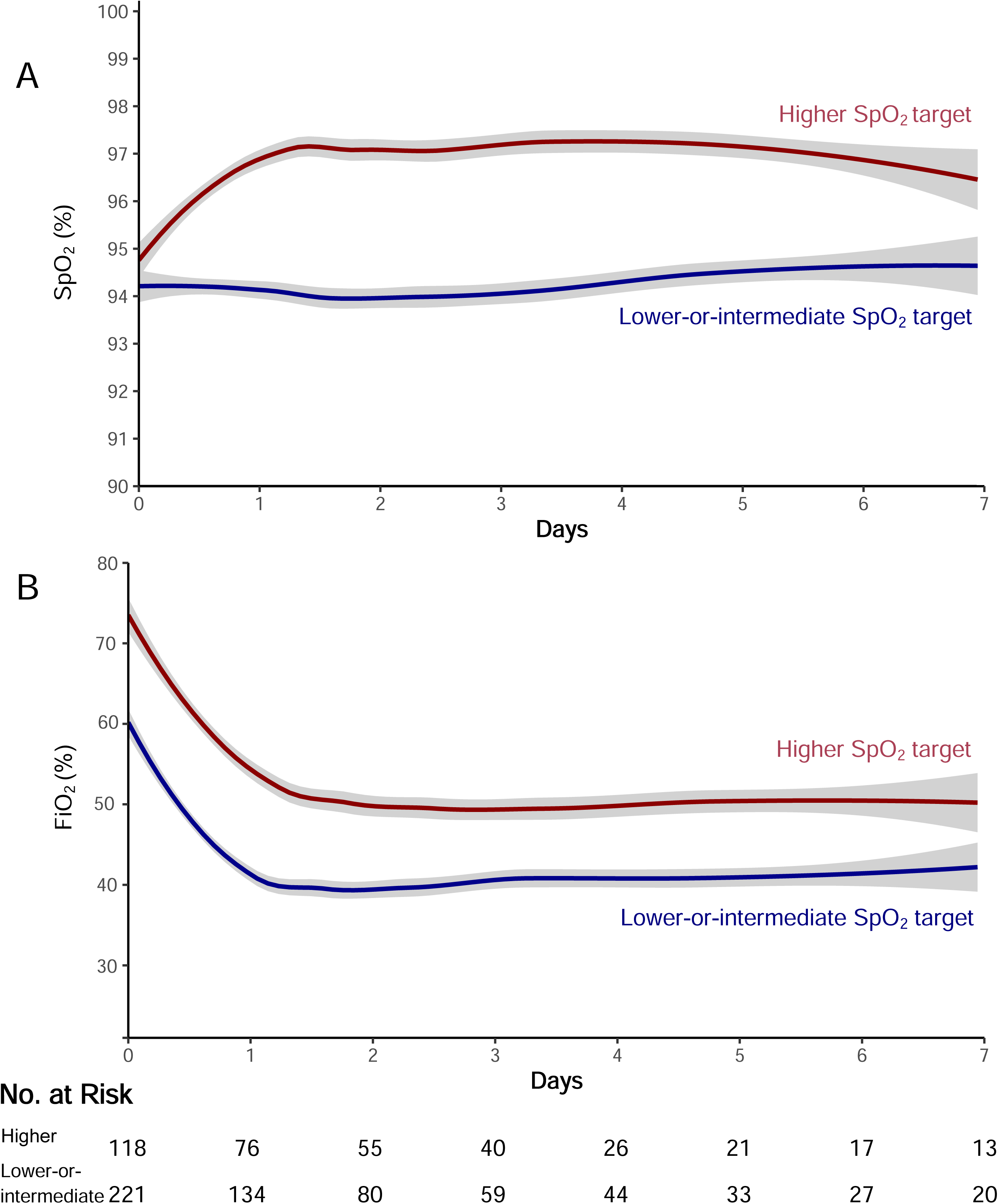
SpO_2_ and FiO_2_ values in each group. Shown are the mean values (colored lines) and 95% confidence intervals (gray shading) for the hourly mean oxygen saturation as measured by pulse oximetry (SpO_2_) (Panel A) and the fraction of inspired oxygen (FiO_2_) (Panel B) from enrollment to study day 7. The data were censored at the time that invasive mechanical ventilation was discontinued. SpO_2_ and FiO_2_ values were obtained approximately every 1 minute, and hourly means were calculated by averaging all measurements obtained during the hour. The number of patients who were alive and receiving invasive mechanical ventilation in each group on each day is shown on the bottom panel.

A single mean SpO_2_ value was calculated for each patient; the median of these values was 95% (IQR 93 to 97%) in the combined lower-or-intermediate target group, and 97% (IQR 96 to 99%) in the higher target group. Similarly, values for FiO_2_ administered were lower in patients assigned to the lower-or-intermediate SpO_2_ targets than those assigned to the higher SpO_2_ target (Figure 2).

A favorable neurologic outcome at hospital discharge occurred in 50 patients (22.6%) assigned to the lower-or-intermediate SpO_2_ targets and 15 patients (12.7%) assigned to the higher SpO_2_ target (absolute risk difference, 9.9 percentage points; 95% CI, 1.8 to 18.1; P=0.03) (Figure 3). Results were similar in the adjusted analysis (adjusted odds ratio, 2.24; 95% CI 1.11 to 4.53; P=0.02) and in the analysis treating Cerebral Performance Category scale as an ordinal outcome ranging from 1 (good neurologic outcome) to 5 (death), (odds ratio, 1.63; 95% CI, 0.99 to 2.68; P=0.05).

**Figure 3.**
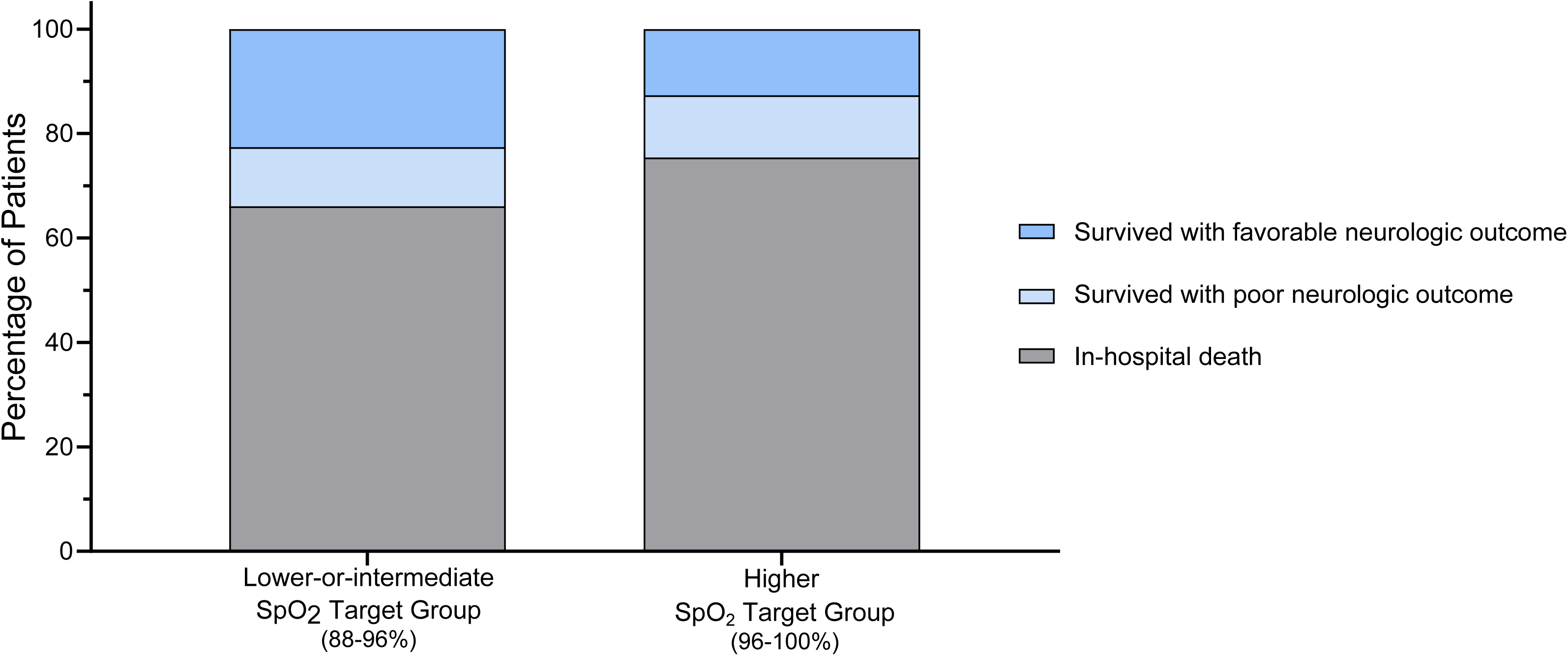
Results of the primary outcome (favorable neurologic outcome) and secondary outcome (in-hospital death). A favorable neurologic outcome at hospital discharge was defined as a Cerebral Performance Category of 1 (good cerebral performance) or 2 (moderate cerebral disability). Alive with poor neurologic outcome was defined as Cerebral Performance Category of 3 (severe cerebral disability) or 4 (coma or vegetative state). Category 5 was in-hospital death. A favorable neurological outcome occurred in 50 patients (22.6%) assigned to a lower-or-intermediate SpO_2_ target and 15 patients (12.7%) assigned to a higher SpO_2_ target (P=0.03).

At 28 days after enrollment, 146 patients (66.1%) assigned to the lower-or-intermediate SpO_2_ targets and 89 patients (75.4%) assigned to the higher SpO_2_ target died prior to hospital discharge (absolute risk difference,-9.4 percentage points; 95% CI,-19.3 to 0.6; P=0.08).

Additional outcomes are presented in Table 2.

**Table 2.** Outcomes.

| Outcomes | Lower-or-intermediate<br>SpO<sub>2</sub> Target<br>(N=221) | Higher<br>SpO<sub>2</sub> Target<br>(N=118) | Risk Difference (RD),<br>Odds Ratio (OR), or<br>Adjusted Odds Ratio<br>(aOR) and (95%CI) | P |
| --- | --- | --- | --- | --- |
| **Primary Outcome** |  |  |  |  |
| Favorable Neurologic Outcome – no (%) <sup>a</sup> | 50 (22.6) | 15 (12.7) | RD 9.9 (1.8 to 18.1)<br>OR 2.00 (1.07 to 3.76) | 0.03 |
| **Additional Analyses of Neurologic Outcome** |  |  |  |  |
| Favorable Neurologic Outcome – no (%), adjusted comparison <sup>b</sup> | 50 (22.6) | 15 (12.7) | aOR 2.24 (1.11 to 4.53) | 0.02 |
| Cerebral Performance Category – no (%), ordinal <sup>c</sup> |  |  | OR 1.63 (0.99 to 2.68) | 0.05 |
| Category 1 | 24 (10.9) | 8 (6.8) |  |  |
| Category 2 | 26 (11.8) | 7 (5.9) |  |  |
| Category 3 | 20 (9.1) | 11 (9.3) |  |  |
| Category 4 | 5 (2.3) | 3 (2.5) |  |  |
| **Secondary Outcome:** |  |  |  |  |
| In-Hospital Death – no. (%) | 146 (66.1) | 89 (75.4) | RD -9.4 (-19.3 to 0.6) | 0.08 |
| **Exploratory Outcomes:** |  |  |  |  |
| Discharge Home – no. (%) <sup>d</sup> | 44 (19.9) | 14 (11.9) | RD 8.0 (0.18 to 15.9) | 0.06 |
| Ventilator- Free Days – median (IQR) <sup>e</sup> | 0 (0 to 22) | 0 (0 to 0) | OR 1.62 (0.98 to 2.68) | 0.06 |
| Mean +/- SD | 7.6 +/- 11.2 | 5.3 +/- 9.9 |  |  |

IQR; interquartile range, SD; standard deviation
a The primary outcome, favorable neurologic outcome, was defined as a Cerebral Performance Category of 1 (good cerebral performance) or 2 (moderate cerebral disability) at the time of hospital discharge.<sup>26</sup> The interrater agreement of favorable versus unfavorable outcome between the first two independent reviewers resulted in a Kappa of 0.66. A chi-squared test was used for between group testing of the primary outcome.
b The adjusted analysis of the primary outcome used a logistic regression model that included the following prespecified co-variables: age, sex, initial shockable rhythm, witnessed arrest, cause of arrest, and time from study start (in days). An odds ratio of greater than 1.0 indicates higher

## DISCUSSION

Among adult patients receiving invasive mechanical ventilation after cardiac arrest in this secondary analysis of a randomized trial, use of a lower-or-intermediate SpO_2_ target (SpO2 88-96%) was associated with a higher incidence of a favorable neurologic outcome at hospital discharge compared to use of a higher SpO_2_ target (SpO2 96-100%).

Preclinical data suggest that a lower oxygenation target may be associated with improved neurologic outcomes in survivors of cardiac arrest, but recent clinical research has generated conflicting evidence.^6–9,30^ The EXACT trial compared oxygen titration to achieve an SpO_2_ of 90-94% versus an SpO_2_ of 98-100% among patients in the prehospital or emergency department setting who had experienced an out-of-hospital cardiac arrest and found lower survival in the 90-94% group (absolute risk difference-9.6%; 95% CI-18.9 to-0.2).^16^ This trial suggests that early oxygen titration, often before SpO_2_ can be reliably measured, may lead to hypoxemia and worse clinical outcomes. The BOX trial compared a restrictive oxygenation target (PaO_2_ of 68 to 75mmHg) versus a liberal oxygenation target (PaO_2_ of 98-105mmHg) in comatose survivors of out-of-hospital cardiac arrest and found no difference in a composite of death or hospital discharge with severe disability or coma (hazard ratio, 0.95; 95% confidence interval, 0.75 to 1.21; P=0.69).^14^ Notably, both the restrictive and liberal PaO_2_ targets in the BOX trial correlated with SpO_2_ values within the low-or-intermediate target ranges of the PILOT trial; few patients experienced severe hyperoxemia. In the ICU-ROX trial, the suspected hypoxic-ischemic encephalopathy subgroup demonstrated significantly lower death in the conservative-oxygen group (SpO_2_ target of less than 97%) than the usual care group (relative risk, 0.73; 95% CI, 0.54 to 0.99) suggesting a potential benefit in this population.

Most prior trials of oxygenation targets after cardiac arrest are limited to patients outside of the United States and those with an out-of-hospital cardiac arrest and an initial shockable rhythm. Little evidence is available to inform care for the 300,000 patients each year who experience an in-hospital cardiac arrest and patients with a non-shockable initial rhythm – the patient populations largely represented in the current study.^14,16^ Our results support the use of a lower SpO_2_ target to avoid hyperoxemia in patients after cardiac arrest may be beneficial.^12,13^ Multiple ongoing randomized trials may help further address oxygenation targets in this population (NCT05029167 and NCT03138005).^31^

This secondary analysis has several strengths. The PILOT trial collected SpO_2_ and FiO_2_ values every minute, which allows for a highly granular assessment of the oxygenation and oxygen therapy experienced by patients in this analysis. Further, this trial enrolled patients at the first receipt of mechanical ventilation in the study emergency department or intensive care unit, capturing the early period in which the harmful effects of hyperoxia are thought to be most pronounced.^32^ Most patients (58.7%) in this analysis experienced an in-hospital cardiac arrest, which is a patient group not well represented by most previous trials evaluating oxygenation targets after cardiac arrest. Finally, this analysis assessed functional outcomes such as the Cerebral Performance Category scale (analyzed as both a binary and ordinal outcome) and discharge destination.

This study also has several limitations. The pragmatic trial and this secondary analysis used only data available from the electronic medical record, which limits the data available for assessment of baseline characteristics and outcomes available for analysis. The trial enrolled patients from a medical intensive care unit; resulting in a high proportion of patients with in-hospital cardiac arrest and without shockable rhythms. A study of this population fills an important knowledge gap but also limits our ability to compare these findings to prior trials.

Sample size limited our ability to definitively evaluate mortality as an outcome.

## INTERPRETATION

Among adults receiving mechanical ventilation after cardiac arrest in a randomized clinical trial of SpO_2_ targets, survival with a favorable neurologic outcome occurred more often with use of a lower-or-intermediate SpO_2_ target (88-96%), compared to use of a higher SpO_2_ target (96-100%). Additional randomized clinical trials are needed to confirm these findings, particularly among patients after cardiac arrest without an initial shockable rhythm.

## Supporting information

Details of the Model Used for Adjusted Outcome

## Data Availability

All data produced in the present study are available upon reasonable request to the authors.
Note this manuscript is currently under peer review at CHEST.

