## Supplementary material for "Effect of Oxygen Saturation Targets on Neurologic Outcomes after Cardiac Arrest: A Secondary Analysis of the PILOT Trial": Details of the Model Used for Adjusted Outcome

Supplementary Appendix

### **Details of the Logistic Regression Model Used for Adjusted Outcomes**

To account for relevant covariates, we fit a logistic regression model with the primary outcome as the dependent variable, and independent variables for study group (lower-or-intermediate target vs higher target) and the following pre-specified baseline covariates: age (continuous), sex (binary), initial shockable rhythm (binary), witnessed arrest (categorical), cause of arrest (categorical), and time from study start (continuous) using a complete case analysis.

| **Adjusted primary analysis** | | | |
| --- | --- | --- | --- |
| **Variable (n=285)** | **Odds Ratio** | **95% CI** | **P-value** |
| Lower-or-intermediate SpO_2_ Target | 2.24 | (1.11, 4.52) | 0.02 |
| Age | 0.99 | (0.97, 1.01) | 0.24 |
| Sex (female) | 1.56 | (0.82, 2.98) | 0.18 |
| Initial Shockable Rhythm | 0.73 | (0.24, 2.20) | 0.54 |
| Witnessed Arrest |  |  |  |
| Bystander | 9.47 | (0.98-91.71) | 0.05 |
| Medical Team | 43.27 | (4.7, 398.93) | 0.001 |
| Unwitnessed | referent |  |  |
| Cause of Arrest |  |  |  |
| Respiratory Failure | 0.35 | (0.10, 1.30) | 0.12 |
| Shock | 0.24 | (0.06, 1.00) | 0.05 |
| Cardiac | 0.44 | (0.09, 2.16) | 0.31 |
| Other Medical | 0.17 | (0.05, 0.68) | 0.01 |
| Non-medical | referent |  |  |
| Time from Study Start (Days) | 1.00 | (0.99, 1.00) | 0.08 |
